# Depression and associated factors among School Age Children with Sickle Cell Disease at Jinja Regional Referral Hospital, Eastern Uganda

**DOI:** 10.64898/2026.01.12.26343983

**Authors:** Maimun Abdiaziz ibrahim, Benedict Akimana, Yahya Abdulkadir Yusuf, Aja Patrick Maduabuchi, Nakawuki Madrine, Sadia Mahad Mohamed, Hafsa Ahmed Ali, Fathi Ali Araye, Abdiaziz Yusuf Haji Ali, Muse Mohamed Mohamoud, Abshir Mohamud Hirsi, Godfrey Zari Rukundo

**Affiliations:** Department of Psychiatry, Faculty of clinical Medicine and Dentistry, Kampala International University, Ishaka, Uganda; Department of Nutritional Nutraceuticals, Kampala International University, Ishaka, Uganda; Department of Psychiatry and behavioral Neurosciences, McMaster University, Hamilton, Ontario, Canada; Department of Internal Medicine, Faculty of clinical Medicine and Dentistry, Kampala International University, Ishaka, Uganda; Department of Obstetrics and Gynecology, Faculty of clinical Medicine and Dentistry, Kampala International University, Ishaka, Uganda; Department of Psychiatry, Belgorod State University, Russia; Department of Public Health, Mount Kenya University, Kenya

## Abstract

**Introduction:** Sickle cell disease is a chronic hematologic disorder associated with significant physical and psychological challenges, including depression. Children with SCD experience recurrent pain crises, hospitalizations, and social limitations, which can contribute to mental health issues. This study aimed to determine the prevalence of depression and associated factors among school-age children with SCD at Jinja Regional Referral Hospital (JRRH), Eastern Uganda.

**Methods:** A cross-sectional study was conducted among 200 randomly selected children aged 6–12 years receiving care at JRRH. Depression was assessed using the Children’s Depression Inventory (CDI). We assessed the association between depression and several factors including sociodemographic characteristics, clinical factors and health related quality of life. Multivariate logistic regression was used to identify factors that were statistically significantly associated with depression at a 95% confidence interval.

**Results:** Majority of the participants 55.5% (111/200) were female, 49.5% (99/200) were in pre-primary with a mean age of 6.7 (SD±1.4) years -The prevalence of depression among children with SCD was 43% (95% CI= 40-46%). Among these, 27.9% had mild depression, 58.1% had moderate depression, and 14.0% had severe depression. Factors significantly associated with depression included lack of assured income among caregivers (AOR=3.67, 95% CI=1.35-7.56, p=0.001), having more than one sibling with SCD (AOR=2.54, 95% CI=1.45-4.72, p=0.02), frequent hospital admissions (AOR=2.12, 95% CI=1.56-4.39, p=0.01), and frequent pain crises (AOR=2.10, 95% CI=1.56-4.67, p<0.001).

**Conclusion:** Depression is prevalent among children with SCD at JRRH, with socio-economic status of the caregiver, number of siblings with SCD, health facility admission frequency and frequent pain crises playing significant roles. Improving access to financial and social support for caregivers and ensuring adequate pain management are recommended.

## Introduction

Globally, depression is a significant comorbidity among children with sickle cell disease (SCD), with prevalence estimates ranging from 13% in Saudi Arabia[1] to 86.4% DRC [4] [2]. In other countries, the prevalence is in between. For example, in the United States, it is 40% [3]; Saudi Arabia 13% [4], South Africa,35.2% [10],. The psychological burden of living with a chronic illness, frequent hospitalizations, and chronic pain significantly impact the mental well-being of affected children and their families, including their health-related quality of life [6] [7]. Despite growing awareness, mental health services for children with SCD remain inadequate in many developing countries including those in Africa [9].

In sub-Saharan Africa, where two-thirds of children with SCD are born annually, the burden of disease is exacerbated by limited healthcare access and inadequate mental health support [10]. Cultural perceptions of mental health and coping mechanisms may influence reported prevalence. The limited availability of psychological support services further compounds the burden of depression in affected children and adolescents and their families [12].

In Uganda, there is a significant knowledge gap regarding the prevalence of depression and associated factors among children with SCD [13] especially those of school-age [10]. The absence of data makes it challenging to implement targeted interventions to improve mental health outcomes and overall quality of life. Given the high prevalence of the sickle cell trait of 17.5% in eastern Uganda [14], there is a critical need to assess the mental health burden in children with SCD. This study aimed to determine the prevalence of depression and associated factors among school-age children with SCD at Jinja Regional Referral Hospital in eastern Uganda.

## Methods and materials

### Study design and setting

This was a cross-sectional study in which we assessed the prevalence of depression and associated factors among children with SCD using questionnaires. The study site was Jinja Regional Referral Hospital which is located in Jinja city. It serves the districts of Bugiri, Iganga, Jinja, Kaliro, Kamuli, Luuka, Mayuge, Namayingo, Kayunga and Buikwe. The hospital is located approximately 84 km east of Mulago National Referral Hospital which is in Kampala the capital city. The psychiatry department in the hospital consist of psychiatrists, Residents (senior house officers), psychiatric clinical officers and psychiatric nurses that are involved in the care of the patients with mental disorders. The psychiatry department manages patient with a wide range of psychiatric conditions including depression. The hospital also has a department of pediatrics that is primarily responsible for the management and follow up of children with sickle cell disease.

### Study participants

This study was conducted among children aged 6-12 years presenting for management of SCD at Jinja Regional Referral Hospital. Written informed consent was obtained from parents or guardians of the participants and assent was obtained from children aged 7 or above. Participants whose Glasgow comma scale was less than 15 were excluded.

### Sampling

We used systematic random sampling to select 200 of the 585 children with SCD at the facility. Averagely, 50 SCD children visit Jinja Regional Referral Hospital on a daily. The sampling interval was calculated as 4 by dividing 200 by 20. The first participant was randomly selected from the first 4, and subsequent participants were selected by adding the interval of 4 to the previous selection. Study participants were always recruited 3 days in a week; i.e. Monday, Wednesday and Friday.

### Study variables

The outcome variable was depression. This was assessed using the Children’s Depression Inventory (CDI), a 27-item self-report tool that measures symptoms such as negative mood, interpersonal problems, ineffectiveness, low self-esteem, and anhedonia. Each item was scored from 0 to 2, with a total score ranging from 0 to 54. A score of 19 or higher indicated clinical depression. Independent variables included socio-demographic characteristics and clinical factors.

### Data collection procedures

Caregivers (parents or guardians) were informed about the study and asked to provide written informed consent, while children provided assent when appropriate. A structured questionnaire was administered to gather socio-demographic and clinical information. Depression was assessed using the CDI. The data collection process started on September 24^th^, 2024, and ended on December 24^th^, 2024. The data collection process ensured privacy and confidentiality, with electronic records stored in password-protected files and paper records kept in locked cabinets. Standard operating procedures for Ebola prevention were adhered to throughout the study.

### Data collection instruments

The data collection instruments used included a structured questionnaire, the CDI, and PedsQL. The structured questionnaire gathered socio-demographic and clinical information about the children and their caregivers, such as age, sex, education level, income, and clinical history related to SCD. The CDI, a 27-item self-report tool, was employed to assess depression levels in the children, measuring symptoms like negative mood, interpersonal problems, and low self-esteem.

### Data quality control

Quality control measures were implemented to ensure the accuracy, reliability, and validity of the study findings. The data collection tools, including the structured questionnaire, Children’s Depression Inventory (CDI), and Pediatric Quality of Life Inventory (PedsQL), were pretested to assess their effectiveness in capturing relevant data. Research assistants were trained on proper data collection procedures to maintain consistency. Daily data checks and verification were conducted by the principal investigator to identify and correct errors. To ensure data integrity, all collected information was double-checked and securely stored, with electronic records protected by passwords and paper records kept in locked cabinets. Additionally, the data collection tools were translated into the locally spoken language (***Lusoga***) for participants who did not speak English to ensure comprehension and minimize bias. These measures enhanced the credibility and reliability of the study outcomes.

### Data management and analysis

Data was entered into Microsoft Excel, cleaned, and then exported to SPSS version 22 for analysis. For the prevalence of depression, percentages and 95% confidence intervals (CIs) were calculated. Bivariate analysis using binary logistic regression identified potential factors associated with depression, and variables with p ≤ 0.2 were further analyzed using multivariate logistic regression to determine independent predictors (p ≤ 0.05 was considered statistically significant). Results were summarized in tables and figures for interpretation.

## Results

### Socio-demographic and clinical variable

Majority of participants 52.5% (105/200) were aged between 6 and 7 years, 55.5% (111/200) were female, and 49.5% (99/200) were in preprimary. Source of social support was mainly from mothers 40.0% (80/200). Furthermore, 44.5% (89/200) of the caregivers were aged less than 20 years, stopped in pre-primary 49.5% (99/200) with majority 60.5% (121/200) not having an assured income. Most caretakers 61.5% (123/200) reported more than 3 siblings with SCD and 1-3 admissions the previous year 49.0% (98/200). Also 54.0% (108/200) reported to have had 1 transfusion, 56.5% (113/200) had Hemoglobin level of 5-8.9 g/dL (table 1).

**Table 1:** Characteristics of participants and their caregivers.

| Variables | Frequencies (N=200) | Total (%) |
| --- | --- | --- |
| Age (years) |  |  |
| 6-7 | 105 | 52.5 |
| 8-9 | 53 | 26.5 |
| 10-12 | 42 | 21.0 |
| Sex |  |  |
| Male | 89 | 44.5 |
| Female | 111 | 55.5 |
| Level of education |  |  |
| Pre-primary | 99 | 49.5 |
| Primary | 71 | 35.5 |
| Not in school | 30 | 15.0 |
| Source of social support |  |  |
| Father | 47 | 23.5 |
| Mother | 80 | 40.0 |
| Grandparents | 52 | 26.0 |
| Others | 21 | 20.5 |
| Age of caregiver (years) |  |  |
| <20 | 89 | 44.5 |
| 20-29 | 39 | 19.5 |
| 30-39 | 25 | 12.5 |
| 40-49 | 14 | 7.0 |
| 50 and plus | 33 | 16.5 |
| Educational level of caregiver |  |  |
| Pre-primary | 99 | 49.5 |
| Primary | 71 | 35.5 |
| Post-secondary | 30 | 15.0 |
| Monthly income of caretaker |  |  |
| No assured income | 121 | 60.5 |
| Less than 200,000 | 37 | 18.5 |
| 200,000-500000 | 24 | 12.0 |
| 500,000-1,000000 | 10 | 5.0 |
| >1000000 | 8 | 4.0 |
| Number of siblings with SCD |  |  |
| ≤1 | 77 | 38.5 |
| >1 | 123 | 61.5 |
| Duration of symptoms yearly |  |  |
| 0-3 times | 31 | 15.5 |
| 4-7 times) | 102 | 51.0 |
| 8-10 times | 67 | 33.5 |
| Number of admissions last year |  |  |
| 0-3 times | 98 | 49.0 |
| 4-7 times | 41 | 20.5 |
| 8-10 times | 61 | 30.5 |
| Number of pain crisis last year |  |  |
| 0-3 times | 112 | 56.0 |
| 4-7 times | 53 | 26.5 |
| 8-10 times | 35 | 17.5 |
| Number of transfusions |  |  |
| 1 | 108 | 54.0 |
| 2 | 60 | 30.0 |
| >3 | 32 | 16.0 |
| Current Hb |  |  |
| < 5.0 | 73 | 36.5 |
| 5-8.9 | 113 | 56.5 |
| >9.0 | 14 | 7.0 |

**Table 2:** Multivariate analysis of the factors associated with depression among children with SCD.

| Variables | COR (95%CL) | P-value | AOR (95%CI) | P-value |
| --- | --- | --- | --- | --- |
| Age (years) |  |  |  |  |
| 6-7 | Ref | - | - | - |
| 8-9 | 1.20(0.76-2.67) | 0.38 | 1.16(0.23-4.34) | 0.54 |
| 10-12 | 1.72(0.87-2.50) | 0.14 | 1.39(0.31-2.56) | 0.37 |
| Sex |  |  |  |  |
| Male | 1.55 (0.88-2.74) | 0.13 | 1.37(0.45-3.92) | 0.18 |
| Female | Ref | - | - | - |
| Level of education |  |  |  |  |
| Pre-primary | 1.60(0.79-3.70) | 0.73 | 1.39(0.45-2.81) | 0.27 |
| Primary | Ref |  |  |  |
| Not in school | 1.84(0.93-1.96) | 0.17 | 1.74(0.49-2.75) | 0.74 |
| Source of social support |  |  |  |  |
| Father | 1.04(0.91-1.74) | 0.098 | 1.12(0.56-2.96) | 0.48 |
| Mother | Ref | - | - | - |
| Grandparent | 0.57(0.04-1.07) | 0.067 | 0.76(0.16-2.49) | 0.13 |
| Other | 2.57(1.03-3.72) | 0.006 | 2.16(0.87-2.87) | 0.09 |
| Age of caregiver (years) |  |  |  |  |
| <20 | 2.86 (0.74-4.96) | 0.13 | 1.87(0.76-4.37) | 0.47 |
| 20-29 | 3.14 (0.61-5.05) | 0.12 | 1.56(0.56-3.78) | 0.78 |
| 30-39 | 1.72 (0.37-6.61) | 0.48 | 1.33(0.87-4.57) | 0.51 |
| 40-49 | Ref | - | - | - |
| 50 and plus | 4.40 (1.03-5.74) | 0.045 | 1.78(0.54-3.69) | 0.39 |
| Educational level of caregiver |  |  |  |  |
| Pre-primary | 1.60(0.43-2.38) | 0.17 | 1.43(0.50-2.68) | 0.54 |
| Primary | Ref | - | - | - |
| Post-secondary | 1.84(0.39-2.60) | 0.73 | 1.34(0.65-4.81) | 0.73 |
| Monthly income of caretaker |  |  |  |  |
| No assured income | 4.80(1.67-6.56) | 0.03* | 3.67(1.35-7.56) | 0.001* |
| Less than 200,000 | 3.17(0.98-5.42) | 0.08* | 2.45(0.78-4.70) | 0.10 |
| 200,000-500000 | 1.24(0.54-2.18) | 0.81 | 1.76(0.54-3.46) | 0.23 |
| 500,000-1,000000 | 0.75(0.16-1.99) | 0.54 | 1.06(0.23-1.87) | 0.30 |
| >1000000 | Ref | - | - | - |
| Number of siblings with SCD |  |  |  |  |
| ≤1 | Ref | - | - | - |
| >1 | 1.56(1.06-3.17) | 0.03 | 2.54(1.45-4.72) | 0.02* |
| Duration of symptoms yearly |  |  |  |  |
| 1-3 times | Ref | - | - | - |
| 4-7 times | 1.27(0.58-2.76) | 0.215 | 1.31(0.43-1.89) | 0.37 |
| 8-10 times | 1.71(1.05-3.79) | 0.04 | 1.04(0.54-6.10) | 0.69 |
| Number of admissions last year |  |  |  |  |
| 0-3 times | Ref | - | - | - |
| 4-7 times | 1.46(0.65-3.61) | 0.09* | 1.56(0.67-3.24) | 0.13 |
| 8-10 times | 4.23(1.57-7.69) | 0.02* | 2.12(1.56-4.39) | 0.01* |
| Number of pain crisis last year |  |  |  |  |
| 0-3 times | Ref | - | - | - |
| 4-7 times | 1.18(0.83-2.07) | 0.24 | 1.87(0.43-3.91) | 0.54 |
| 8-10 times | 1.83(1.21-4.69) | 0.015 | 2.10(1.56-4.67) | <0.001* |
| Current Hb |  |  |  |  |
| < 5.0 | 1.66(0.78-2.70) | 0.19 | 1.13(0.43-2.67) | 0.40 |
| 5-8.9 | 1.24(0.54-5.78) | 0.96 | 1.05(0.05-5.80) | 0.91 |
| >9.0 | Ref | - | - | - |

Statistically significant, P<0.05

### Prevalence of depression among school age children with SCD

In general, the prevalence of depression in SCD children was 43% (95% CI= 40-46%). Of these, over half 58.1% (50/86) had moderate form of depression and 27.9% (24/86) had mild form. About 14.0% (12/86) had severe form of depression (Figure 4).

### Factors associated with depression among children with SCD

At bivariate regression analysis, age of the child, sex, education level, source of social support, age of caregiver, education level of caregiver, monthly income of caregiver, number of siblings with SCD, annual duration of symptoms, number of hospital admissions in the previous year, number of pain crisis last year and current Hb levels were significantly associated with the outcome variable.

At multivariate regression analysis, caretakers with no assured income [AOR=3.67, CI=1.35-7.56, P=0.001] were 3.67 times more likely to have depressed children compared to those who earned more than 1 million at a 95% confidence interval. Children with >1 sibling with SCD [AOR=2.54, CI=1.45-4.72, P=0.02] were 2.54 times more likely to be depressed as compared to those with ≤1 sibling with SCD. Those who were admitted 8-10 times in a year [AOR=2.12, CI=1.56-4.39, P=0.01] were 2.12 times more likely to be depressed as compared to those who were admitted 1-3 times in a year. Furthermore, those who experienced pain crisis 8-10 times last year [AOR=2.10, CI=1.56-4.67, P<0.001] were 2.10 more likely to be depressed as compared to those who experienced pain crisis 1-3 times last year (Table 3).

## Discussion

This study aimed to determine the prevalence of depression and associated factors among children SCD at Jinja regional referral hospital, Eastern Uganda. We found a prevalence of depression at 43%. The associated factors depression was: parent’s monthly income, number of siblings with SCD, previous year’s number of admissions and previous year’s pain crises.

### Prevalence of Depression among Children with Sickle Cell Disease (SCD)

The prevalence of depression among children with sickle cell disease (SCD) in this study was found to be 43%, with the majority experiencing moderate depression (58.1%), followed by mild (27.9%) and severe depression (14.0%). This prevalence is comparable to studies conducted in other regions, though variations exist. For instance, a study in South Africa reported a prevalence of 35.2% [11], which is slightly lower than our findings. The lower prevalence in the latter study compared to ours could be attributed to more investments in mental health issues especially among children by parents and caregivers. In contrast, a study in Congo found a much higher prevalence of 86.4% [5], indicating significant differences in mental health burden across populations. The disparity may be attributed to differences in healthcare access, socio-economic conditions, and cultural perceptions of mental health.

In high-income countries such as the United States, the prevalence of depression among children with SCD has been reported at approximately 40–50% [3], similar to the findings in this study. The relatively high prevalence of depression among children with SCD in this setting can be linked to factors such as chronic pain, frequent hospitalizations, social stigma, and limited mental health support. The psychological burden of living with a chronic condition, coupled with socio-economic constraints and reduced quality of life, significantly contributes to the high rates of depression observed in these children.

### Factors Associated with Depression Among Children with SCD

In this study, children whose caregivers had no assured income were 3.67 times more likely to have depression than those from financially stable households. This finding aligns with studies in Nigeria and Saudi Arabia, which found that lower household income and financial instability increased the likelihood of depressive symptoms in children with SCD [7, 8]. The financial burden of managing SCD, including medication costs, transportation to hospitals, and dietary needs, may contribute to stress in caregivers and emotional distress in children. However, some studies have found no significant association between caregiver income and depression. For example, a study in the United States [15] did not find a direct link between socioeconomic status and depression among children with SCD. This discrepancy may be due to differences in healthcare systems. In high-income settings like the United States, financial assistance programs, health insurance, and subsidized medical care may buffer the effects of low income, reducing its impact on mental health. In contrast, in low-resource settings like Uganda, where access to free or subsidized healthcare is limited, financial constraints may directly affect a child’s ability to receive timely medical attention, contributing to higher rates of depression.

Our study found that children with more than one sibling affected by SCD were 2.54 times more likely to be depressed than those with fewer or no affected siblings. This aligns with findings from a study in Saudi Arabia, where larger family size and multiple cases of SCD within a household were associated with higher depression rates [16]. The explanation could be that multiple cases of SCD in a household place an increased financial and emotional burden on caregivers, limiting their ability to provide individualized psychological support to each child. Additionally, children in such households may experience a sense of neglect and perceive themselves as burdens, which could contribute to depressive symptoms. However, some studies, such as one conducted in Brazil [6], did not find a significant association between the number of siblings with SCD and depression. This difference could be attributed to variations in family dynamics, cultural perceptions of chronic illness, and the availability of social support systems. In some cultures, larger families may offer better emotional and psychological support through sibling bonding, reducing the risk of depression.

In this study, children who had been admitted to the hospital 8–10 times in the past year were 2.12 times more likely to be depressed than those with fewer admissions. This finding is consistent with studies from Nigeria and Saudi Arabia, where frequent hospital visits were linked to higher levels of depression in children with SCD [8, 17]. The repeated disruption of daily life, school absenteeism, and exposure to painful medical procedures could contribute to a heightened sense of distress and anxiety. However, a study in South Africa [11] found no significant association between hospital admissions and depression. A possible explanation for this discrepancy is that in some settings, children who are frequently hospitalized may receive better medical attention, pain management, and psychological support, mitigating the emotional toll of hospitalization. Additionally, differences in study methodology, including how depression was assessed and the inclusion criteria for hospitalization, may account for the variation in findings.

Children in our study who experienced frequent pain crises (8–10 times in the past year) were 2.10 times more likely to develop depression than those with fewer episodes. This is in agreement with studies from the United States and South Africa, where increased pain frequency was a strong predictor of depression in children with SCD [11, 18]. Chronic pain is known to impact mental health by limiting physical activity, reducing social interactions, and causing emotional distress. In contrast, some studies, such as one conducted in Nigeria [19], did not find a significant association between pain frequency and depression. One possible explanation is that pain perception and coping mechanisms may vary across individuals and cultural contexts. In some communities, strong social support systems and resilience-building interventions may help children manage pain-related distress, reducing the likelihood of depression. Additionally, differences in pain management strategies across healthcare settings could influence the psychological impact of pain crises.

### Clinical implications of the study findings

The high prevalence of depression (43%) among these children indicates the need for routine mental health screening in pediatric SCD clinics. Early identification and intervention for depression can prevent severe psychological distress and improve overall well-being. The significant association between depression and factors such as lack of assured income, multiple siblings with SCD, frequent hospital admissions, and frequent pain crises highlights the importance of a holistic approach to care. This approach should integrate mental health support, effective pain management, and socio-economic assistance for families.

### Strengths and weaknesses of the study

The study used systematic random sampling technique which enhanced the representativeness of the sample hence reducing selection bias and improving the generalizability of the findings.

The study as well had some weakness in terms of its design;

The study’s cross-sectional design limits the ability to establish causality between depression and associated factors. Longitudinal studies would be needed to assess changes over time.

The reliance on self-report tools for assessing depression may introduce response bias due to social desirability or recall issues.

Conducting the study at a single hospital may limit the generalizability of the findings to other regions or settings, as the sample may not fully represent children with SCD in the community.

The exclusion of children with a Glasgow Coma Scale score less than 15 may have omitted a subset of children with severe complications, potentially affecting the overall prevalence and factors associated with depression.

## Conclusion

This study highlights a high prevalence of depression (43%) among children with SCD at JRRH. Several factors, including caregiver income, the number of siblings with SCD, the frequency of hospital admissions, and the frequency of pain crises, were significantly associated with depression. Additionally, children with depression had poorer quality of life, emphasizing the need for integrated mental health interventions in SCD management. These findings highlight the importance of early screening, psychosocial support, and policy measures to improve the well-being of children living with SCD in Uganda.

## Data Availability

All relevant data are within the manuscript and its Supporting Information files.

## Declarations

### Ethical considerations

Ethical approval for the study was obtained from the Research Ethics Committee of Kampala International University (KIU-REC), and permission was granted by Jinja Regional Referral Hospital (JRRH). The study adhered to the ethical principles of voluntary participation, informed consent, confidentiality, and beneficence. Parents or guardians provided written informed consent, while children aged 7 years and above provided assent before participation. Confidentiality was maintained by de-identifying data and storing records securely in password-protected electronic files and locked cabinets. Participants were assured that their participation was voluntary, and they could withdraw at any time without consequences. The study complied with Uganda National Council for Science and Technology (UNCST) guidelines and followed COVID-19 and Ebola prevention protocols, including mask-wearing, hand hygiene, and social distancing where applicable.

### Consent for publication

Not applicable

### Clinical trial Number

Clinical trial number not applicable

### Availability of data and materials

All data are presented within the manuscript

### Authors’ contributions

MAI conceived the original idea of the study. NM, SMM, MMM, AYHA, contributed to the development of the idea into a proposal. MH, HAA, and FAA provided guidance and support to MAI during data collection. BA, YAY, APM and GZR Supervised and participated in data analysis and interpretation of results. MAI wrote the first full draft manuscript, and all authors critically reviewed the manuscript and agreed upon the final version for submission to the journal.

## Acknowledgments

The authors acknowledge the study participants who provided useful information and insights. The research assistants are acknowledged for their dedication during data collection.

## References

1. Sehlo, M.G. and H.Z. Kamfar, Depression and quality of life in children with sickle cell disease: the effect of social support. BMC Psychiatry, 2015. 15(1): p. 78.

2. Leite, A.G.S., et al., Prevalence of clinical manifestations suggestive of depression in patients with sickle cell disease: a review. Jornal Brasileiro de Psiquiatria, 2022. 71: p. 56–62.

3. Hardy, S.J., et al., A randomized controlled trial of working memory training in pediatric sickle cell disease. Journal of Pediatric Psychology, 2021. 46(8): p. 1001–1014.

4. Sehlo, M.G. and H.Z. Kamfar, Depression and quality of life in children with sickle cell disease: the effect of social support. BMC psychiatry, 2015. 15: p. 1–8.

5. Lukoo, R.N., et al., Depression in children suffering from sickle cell anemia. Journal of Pediatric Hematology/Oncology, 2015. 37(1): p. 20–24.

6. de Oliveira, L.A.B., et al., Religiosity, Anxiety, Depression, and suicidal Ideation in Brazilian Patients with sickle cell disease Religiosidade, Ansiedade, Depressão e Ideação suicida em Pacientes Brasileiros com doença falciforme.

7. Zigashane, A.B., et al., Quality of life among patients with Sickle Cell Disease and their parents in Democratic Republic of Congo, qualitative study. 2023.

8. Gemici Karaaslan, H.B., et al., Association of depression and social anxiety symptom scores with disease characteristics in pediatric patients with chronic immune thrombocytopenia: a cross-sectional study. International Journal of Hematology, 2024. 120(3): p. 356–364.

9. Ezenwosu, O.U., et al., Assessment of depression in children and adolescents with sickle cell anemia in a low-resource setting: a comparative study. Pediatric Hematology and oncology, 2023. 40(1): p. 40–50.

10. Atoku, A.C., et al., Psycho-social challenges faced by caretakers of children and adolescents aged 0–19 years with sickle cell disease admitted in a tertiary hospital in Eastern Uganda. Journal of Pediatric Nursing, 2023. 69: p. e21–e31.

11. Adam, S.S., et al., Depression, quality of life, and medical resource utilization in sickle cell disease. Blood advances, 2017. 1(23): p. 1983–1992.

12. Graves, J.K., C. Hodge, and E. Jacob, Depression, anxiety, and quality of life in children and adolescents with sickle cell disease. Pediatric nursing, 2016. 42(3): p. 113.

13. Kambasu, D.M., et al., Health-related quality of life of adolescents with sickle cell disease in sub-Saharan Africa: a cross-sectional study. BMC hematology, 2019. 19: p. 1–9.

14. Alamun, A.I., et al., Psychological Experience of Mothers of Children with Sickle Cell Disease Followed at the Pediatric Department of Bouaké University Teaching Hospital. Open Journal of Pediatrics, 2024. 14(1): p. 149–163.

15. Suliman, M.E., et al., Telomere length and telomere repeat-binding protein in children with sickle cell disease. Pediatric Research, 2022. 91(3): p. 539–544.

16. Weytey, S., The Quality of Life of Children with Sickle Cell Disease (SCD). Current Practices in Sickle Cell Disease, 2024: p. 113.

17. Albayrak, I., et al., Assessment of pain, care burden, depression level, sleep quality, fatigue and quality of life in the mothers of children with cerebral palsy. Journal of Child Health Care, 2019. 23(3): p. 483–494.

18. Prussien, K.V., et al., Cognitive function, coping, and depressive symptoms in children and adolescents with sickle cell disease. Journal of Pediatric Psychology, 2018. 43(5): p. 543–551.

19. Ezenwosu, O., et al., Clinical depression in children and adolescents with sickle cell anaemia: influencing factors in a resource-limited setting. BMC pediatrics, 2021. 21: p. 1–8.

